# Noncanonical WNT activation in human right ventricular heart failure

**DOI:** 10.1101/2020.07.03.20134965

**Authors:** Jonathan J. Edwards, Jeffrey Brandimarto, Dong-Qing Hu, Sunhye Jeong, Nora Yucel, Li Li, Kenneth C. Bedi, Shogo Wada, Danielle Murashige, Hyun Tae Hwang, Mingming Zhao, Kenneth B. Margulies, Daniel Bernstein, Sushma Reddy, Zoltan P. Arany

## Abstract

**Background:** No medical therapies exist to treat right ventricular (RV) remodeling and RV failure (RVF), in large part because molecular pathways that are specifically activated in pathologic human RV remodeling remain poorly defined. Murine models have suggested involvement of Wnt signaling, but this has not been well defined in human RVF.

**Methods:** Using a candidate gene approach, we sought to identify genes specifically expressed in human pathologic RV remodeling by assessing the expression of 28 WNT-related genes in the RVs of three groups: explanted nonfailing donors (NF, n = 29), explanted dilated and ischemic cardiomyopathy, obtained at the time of cardiac transplantation, either with preserved RV function (pRV, n = 78) or with RVF (n = 35).

**Results:** We identified the noncanonical WNT receptor ROR2 as transcriptionally strongly upregulated in RVF compared to pRV and NF (Benjamini-Hochberg adjusted p<0.05). ROR2 protein expression correlated linearly to mRNA expression (R^2^ = 0.41, p = 8.1×10^−18^) among all RVs, and to higher right atrial to pulmonary capillary wedge ratio in RVF (R^2^=0.40, p = 3.0×10^−5^). We compared RVF with high and low ROR2 expression, and found that high ROR2 expression was associated with increased expression of the WNT5A/ROR2/Ca^2+^ responsive protease calpain, cleavage of its target FLNA, and FLNA phosphorylation, another marker of activation downstream of ROR2. ROR2 protein expression as a continuous variable, correlated strongly to expression of calpain (R^2^ = 0.25), total FLNA (R^2^ = 0.67), and FLNA phosphorylation (R^2^ = 0.62, p < 0.05 for all).

**Conclusion:** We demonstrate robust reactivation of a fetal WNT gene program, specifically its noncanonical arm, in human RVF characterized by activation of ROR2/calpain mediated cytoskeleton protein cleavage.

## Introduction

Right ventricular failure (RVF) is independently predictive of morbidity and mortality in diverse disease processes including left ventricular failure (LVF), pulmonary hypertension, and congenital heart disease [1; 2; 3]. There is a large gap, however, in our understanding and management of RVF [4]. Multiple studies demonstrate that standard reverse remodeling agents that unequivocally improve survival for LVF, such as ACE inhibitors and beta-blockers, rarely have impact in RVF, with sometimes even worsened outcomes [3; 5; 6; 7]. Furthermore, markers of left ventricular remodeling poorly predict RV dysfunction [8]. Thus, greater understanding of the fundamental mechanisms that drive RVF, different from LVF, is needed.

Evidence suggests that an RV-specific remodeling transcriptional program contributes to this disparate clinical behavior of the RV and LV [9]. We chose to evaluate *WNT*-related differential gene expression in human RV remodeling for multiple reasons. Canonical Wnt signaling regulates cardiomyocyte proliferation during development, and is critical to second heart field development—which gives rise to the RV—but relatively dispensable for the first heart field— which gives rise to the LV [10; 11]. In parallel, noncanonical Wnt signaling promotes developmental cardiomyocyte maturation, and knockout of downstream genes including *Scrib, Vangl2*, or *Rac1* results in second heart field structural anomalies and altered myocardial patterning and cardiomyocyte cell shape that resemble those seen in pathologic remodeling [12; 13; 14]. In murine models of LVF and *in vitro* cardiomyocyte models, aberrant activation of both canonical and non-canonical Wnt signaling has been connected to cardiomyocyte hypertrophy, fibroblast proliferation and activation, activation of cytoskeleton remodeling, and activation of stress pathways [15; 16; 17; 18; 19]. Unbiased transcriptomics analyses of RVF in mice due to pressure or volume-overload have identified *WNT* signaling as a pathway that is specifically altered in the progression from a compensated to decompensated state ([20; 21]. In humans, a recent study illustrated that higher *WNT5A* serum levels and myocardial expression correlated with worse RV, but not LV, systolic function and with higher likelihood of death or transplant in patients with dilated cardiomyopathy (DCM) [22]. Together, these studies have suggested that embryonic or fetal Wnt expression may be reactivated in RV remodeling, akin to the well-established reactivation of fetal programs in LV remodeling. However these and other studies demonstrating aberrant WNT signaling in RV remodeling have been limited to transcriptomics in murine models or were narrowly designed in humans such that the potential clinical role of WNT signaling in adaptive and pathologic remodeling, with associated preserved function (pRV) and RVF, respectively, remains incompletely defined [20; 21; 22; 23; 24]. Finally, we also focused on WNT signaling because this pathway is dependent on extracellular factors and cell surface receptors, potentially facilitating prognostic or therapeutic avenues that target WNT signaling in RV remodeling.

LVF is an ideal human setting in which to characterize the gene expression signature of adaptive and pathologic RV remodeling for two reasons. LVF is the most common cause of RVF, and LVF causes a range of RV involvement—from pRV to RVF—reflecting different types of adaptive/pathologic RV remodeling or different points in time in disease progression [25; 26]. In this study, we leverage a large collection of human RV tissues from explanted DCM and ischemic (ICM) hearts, stratified with either pRV or RVF, and from nonfailing (NF) hearts from human donors, in order to identify a robust reactivation of the fetal noncanonical WNT receptor ROR2, upregulation of ROR2/Ca^2+^ responsive protease calpain, and increased cleavage of calpain-target cytoskeletal proteins specifically in severe RVF [27; 28; 29]. We propose this pathway as a potential novel therapeutic target of pathologic RV remodeling.

## Materials and Methods

### Human samples

Procurement of all RV myocardial tissue was performed using Gift-of-Life and University of Pennsylvania Institutional Review Board (approval 802781) approved protocols with informed consent provided when appropriate as previously described.[30] RV myocardial samples were retrospectively obtained from the Penn Human Heart Tissue Library collected from May 2005 to April 2018. DCM and ICM hearts were procured at the time of clinical heart transplantation. The RV functional status (pRV vs. RVF) was identified using pretransplant right atrial (RA) pressure and RA to pulmonary capillary wedge pressure ratio (RA:PCWP) as this has ratio is predictive of RVF in the setting of LVF following left ventricular assist device placement in previously published studies [25; 26]. Four cardiomyopathy RV hemodynamic patient groups were identified based on RV hemodynamics:

DCM-RVF and ICM-RVF: RA ≥ 9 and RA:PCWP ≥ 0.63
DCM-pRV and ICM-pRV: RA ≤ 8 and RA:PCWP ≤ 0.37.

Exclusion criteria included prior ventricular assist device, retransplantation, incomplete hemodynamics, or insufficient RV tissue for analysis. Potential clinical confounders to differential gene expression were assessed using chi square analysis test for categorical variables and Mann-Whitney U for continuous variables with pairwise deletion for any missing data. Glomerular filtration rate was calculated using the MDRD equation. All reported p values were adjusted using the Benjamini-Hochberg multiple comparison correction method (adjusted *P* < 0.05 for significance).

Disease groups were compared to a NF cohort, selected from unused Gift-of-Life donor hearts that were deemed unsuitable for transplantation as previously described.[30] We enriched for normal RV function by selecting those with confirmed minimal tricuspid insufficiency, preserved left ventricular function by ejection fraction ≥ 50%, and preserved renal function by creatinine ≤ 1.2.

### RNA expression

Total RNA was extracted from coded frozen human tissue samples using RNeasy Mini Kit (Qiagen Hilden, Germany). RNA samples were diluted to 100 ng/uL by DEPC water using Qubit fluorometer (Thermo Fisher Scientific Waltham, MA). Human cDNA was synthesized using MultiScribe Reverse Transcriptase™ (Thermo Fisher Scientific). Gene expression was quantified using SYBR Green Master Mix RT-PCR. RT-PCR primers (sequences in **Supplemental Table 1**) were validated using primer efficiency and melt curve analyses. Log2fold changes were calculated using the housekeeping genes GAPDH and TBP. Candidate genes were first assessed for differential expression in both DCM and ICM using Kruskal-Wallis test to compare log2fold (adjusted p < 0.05) between NF/pRV/RVF for both the DCM and ICM cohorts. Genes that were differentially expressed for both DCM and ICM were assessed for differential expression between pRV and RVF using Mann Whitney test after combining DCM and ICM groups.

### Protein expression

Genes that demonstrated statistically significant differential expression between pRV and RVF were further assessed using western blot to evaluate differential protein expression. As a preliminary analysis, four representative samples from each group were selected using the lowest and highest RA:RPCW ratio for pRV and RVF, respectively, and the lowest NPPA expression for the NF controls. Protein extraction was performed using NE-PER™ kit (ThermoFisher Scientific) to separate cytoplasmic and nuclear fractions. Equal amount cytoplasmic or nuclear protein according to predicted protein location from each sample was separated by SDS-PAGE. Western blot was performed using monoclonal antibodies to the following targets: CREBBP (Cell Signaling Technology (CST) Danvers, MA, cat D6C5), NFATC2 (Abcam, Cambridge, UK, cat ab2722), ROR2 (CST cat D3B6F), TBP (Abcam, cat ab51841), HDAC2 (Abcam, cat ab32117), and GAPDH (CST, cat D16H11). Given preliminary results demonstrating significant upregulation of ROR2, western blots were performed for all remaining samples. In a subset of RVF samples with either highest (n = 6) or lowest (n = 6) ROR2 expression, we performed total protein extraction using RIPA to explore activation of downstream pathways. Western blot was performed using antibodies to filamin A (FLNA, CST cat 4762), serine-2152 phosphorylated FLNA (CST cat 4761), calpain (CST cat 2556), phosphorylated PAK1 (CST cat 2601), and spectrin (Biolegend, San Diego, CA cat D8B7). Secondary anti-mouse (CST cat 7076) and anti-rabbit (CST cat 7074) were used as indicated. Densitometry was assessed using SuperSignal™ West Femto enhanced chemiluminescent substrate (ThermoFisher Scientific) and ImageStudio (LI-COR Biotechnology, Lincoln, Nebraska).

### Statistics

All data was analyzed using RStudio version 1.1.463. Data are presented as median (interquartile range), count (%), or adjusted R^2^ for correlations. Pairwise deletion was performed for any missing clinical data. Statistical significance was determined using Mann Whitney U or Kruskal-Wallis, where appropriate, for continuous variables, and chi square for categorical variables. All reported p values were adjusted using the Benjamini-Hochberg multiple comparison correction method (adjusted *P* < 0.05 for significance).

## Results

### Patient characteristics

We collected RV myocardial tissue from patients undergoing clinical heart transplantation for LVF due to DCM or ICM and separated them according to the presence of pRV or RVF using preexplant hemodynamic data, as described in the methods. Median time between hemodynamic data collection and explantation was 28.5 days (interquartile range: 13 – 53 days). In total, we identified 47 DCM-pRV, 26 DCM-RVF, 31 ICM-pRV, and 9 ICM-RVF, representing the largest and most clinically diverse study of human RV differential gene expression in LVF to date.

We identified no clinical confounders between DCM-pRV/DCM-RVF, ICM-pRV/ICM-RVF, and combined pRV/RVF groups (**Table 1** and **Supplemental Table 2**), including no differences in gender, age, ethnicity, body surface area, body weight, heart weight, renal function by glomerular filtration rate, diabetes, use of reverse remodeling agents collectively or individually (ACE inhibitor, angiotensin receptor blocker, or β-blocker), use of other cardioactive medications (digoxin, diuretics, calcium channel blockers, or milrinone), lipid lowering medications, thyroid medications, or pacemakers. Also, ICM-RVF hearts were no more likely to have significant right coronary artery disease (≥ 70% stenosis) or history of prior coronary intervention compared to their ICM-pRV counterparts, indicating that any observed transcriptional changes could not be attributable to dichotomous coronary involvement.

**Table 1.** Clinical and demographic characteristics of right ventricular functional status by cardiomyopathy type. ^a^Continuous variables presented as median (interquartile range) and *p* value using two-tailed Mann-Whitney U test. Categorical variables presented as percent and *p* value using Chi square 2×2 contingency tables using pairwise deletion for any missing data. All *p* values are Benjamini-Hochberg corrected, and none were less than 0.05. Abbreviations: angiotensin converting enzyme inhibitor (ACE inhibitor), angiotensin receptor blocker (ARB), coronary artery bypass grafting (CABG), glomerular filtration rate (GFR), and RCA (right coronary artery disease).

| Clinical and demographic characteristics |  |  |  |  |  |  |
| --- | --- | --- | --- | --- | --- | --- |
| Variables | Dilated Cardiomyopathy |  |  | Ischemic Cardiomyopathy |  |  |
|  | RVF<br>n = 26 | pRV<br>n = 47 | Adj p | RVF<br>n = 9 | pRV<br>n = 31 | Adj p |
| Age<sup>a</sup> | 56.5<br>(49, 58) | 52.0<br>(44, 60) | 0.60 | 60.0<br>(56, 62) | 61.0<br>(55, 63) | 1.0 |
| Male | 50% | 72% | 0.60 | 89% | 87% | 1.0 |
| Ethnicity |  |  | 1.0 |  |  | 1.0 |
| Caucasian | 54% | 64% |  | 89% | 84% |  |
| African American | 38% | 30% |  | 0% | 13% |  |
| Other | 8% | 6% |  | 11% | 3% |  |
| Heart weight (grams)<sup>a</sup> | 471<br>(380, 549) | 465<br>(421, 545) | 0.87 | 570<br>(512, 625) | 552<br>(455, 639) | 1.0 |
| Weight (kg)<sup>a</sup> | 76<br>(73, 91) | 80<br>(69, 92) | 1.0 | 93<br>(84, 113) | 82<br>(75, 94) | 0.92 |
| Body surface area (m<sup>2</sup>)<sup>a</sup> | 1.90<br>(1.8, 2.1) | 1.95<br>(1.8, 2.1) | 1.0 | 2.19<br>(2.0, 2.4) | 1.99<br>(1.9, 2.2) | 0.92 |
| GFR<sup>#</sup> | 49.0<br>(43, 65) | 67.3<br>(55, 82) | 0.10 | 55.4<br>(42, 56) | 53.5<br>(44, 70) | 1.0 |
| Diabetes Mellitus | 32% | 28% | 1.0 | 56% | 39% | 1.0 |
| Insulin | 15% | 15% | 1.0 | 44% | 13% | 1.0 |
| Thyroid medication | 31% | 6% | 0.14 | 33% | 16% | 1.0 |
| Pacer | 38% | 21% | 0.60 | 11% | 6% | 1.0 |
| ACE inhibitor | 54% | 51% | 1.0 | 56% | 58% | 1.0 |
| ARB | 31% | 26% | 1.0 | 22% | 16% | 1.0 |
| β-blocker | 81% | 94% | 0.60 | 67% | 87% | 1.0 |
| Any Reverse Remodeling | 100% | 96% | 1.0 | 100% | 94% | 1.0 |
| Calcium Channel Blocker | 8% | 6% | 1.0 | 0% | 0% | 1.0 |
| Digoxin | 50% | 47% | 1.0 | 22% | 45% | 1.0 |
| Diuretic | 100% | 87% | 0.60 | 78% | 90% | 1.0 |
| Lipid lowering | 42% | 49% | 1.0 | 67% | 71% | 1.0 |
| Milrinone | 65% | 72% | 1.0 | 86% | 71% | 1.0 |
| RCA disease | 4% | 0% | N/A | 56% | 74% | 1.0 |
| Prior CABG | 0% | 0% | N/A | 67% | 43% | 1.0 |
| Prior Angioplasty | 4% | 0% | N/A | 56% | 52% | 1.0 |
| Prior Stent | 4% | 0% | N/A | 56% | 52% | 1.0 |

The NF group was similarly well matched across all four cardiomyopathy hemodynamic groups with respect to age, ethnicity, body surface area, and weight, although not for gender (**Supplemental Table 3**). Previous unbiased studies of cardiac DGE have demonstrated few gender-specific differences, and none in WNT-related gene expression, suggesting a low likelihood that gender differences would impact our findings [31].

### WNT Pathway Candidate Gene Analysis

#### RNA expression

In total, we assessed the RV myocardial expression of 28 WNT-related genes including ligands, receptors and co-receptors, inhibitors, and downstream signaling and transcriptional targets that have been either previously implicated in RV remodeling in murine models of RVF or have a known interaction based on literature review (**Supplemental Table 3**). First, we assessed whether these 28 WNT-related genes and two well described heart failure genes—NPPA and NPPB—exhibited differential expression between NF/pRV/RVF separately for both DCM and ICM using Kruskal-Wallis to compare log_2_fold changes.[32] Most WNT-related genes (**Supplemental Table 4**) and both natriuretic peptides demonstrated statistically significant differential expression in at least DCM or ICM, with 12 demonstrating differential expression in both: AXIN2, CREBBP, DAAM2, FZD1, FZD7, NFATC2, NPPA, ROR2, SFRP1, SFRP3, WISP2, and WNT10B. Of these 12, only five genes were found to be differentially expressed between pRV and RVF: CREBBP, NFATC2, NPPA, ROR2, and WISP2 (**Table 2, Figure 1**). To further prioritize these candidate genes, we performed linear regressions comparing RNA expression to RA:PCWP and found that ROR2 (R^2^ = 0.16, p = 3.2 × 10^−5^), CREBBP (R^2^ = 0.03, p = 0.048), and NFATC2 (R^2^ = 0.03, p = 0.048) had modest correlations with the RA:PCWP, while WISP2 did not (R^2^ = 0.01, p = 0.14).

**Table 2.**

| Differential RNA Expression between RVF and pRV |  |  |
| --- | --- | --- |
| Gene | RVF/pRV Fold Expression | Adj p |
| AXIN2 | 1.30 | 0.07 |
| **CREBBP** | **1.64** | **2.63x10<sup>-4</sup>** |
| DAAM2 | 1.15 | 0.46 |
| FZD1 | 1.10 | 0.55 |
| FZD7 | 1.19 | 0.46 |
| **NFATC2** | **1.46** | **0.033** |
| **NPPA** | **2.68** | **0.037** |
| **ROR2** | **1.57** | **0.010** |
| SFRP1 | 0.90 | 0.31 |
| SFRP3 | 1.23 | 0.13 |
| **WISP2** | **1.42** | **0.039** |
| WNT10B | 1.03 | 0.55 |

2 Genes which were differentially expressed in DCM and ICM pRV/RVF compared to NF using 3 Kruskal Wallis were selected for further analysis comparing RVF to pRV expression using 4 Benjamini Hochberg corrected Mann Whitney U test (p <0.05, bolded for significance).

**Figure 1.**
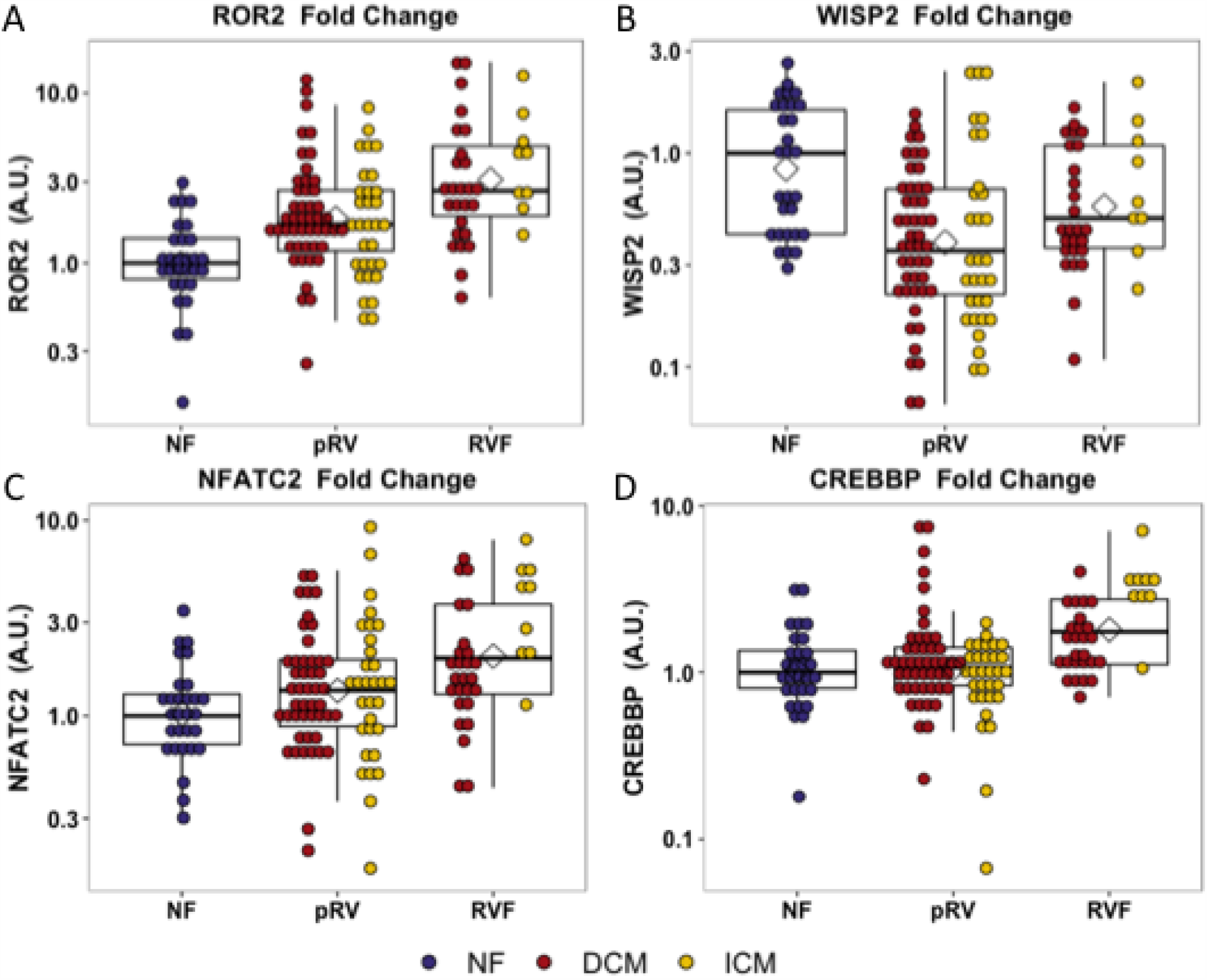
Relative RNA expression of WNT-related genes with differential 1 transcription between pRV and RVF as noted in panels A-D.

#### Protein expression

To assess whether protein expression would correlate with the observed transcriptional upregulation of CREBBP, NFATC2, and ROR2 in RVF, we first performed a preliminary analysis comparing four representative samples from each group representing the hemodynamic extremes for pRV and RVF and the lowest NPPA expression for NF. In this preliminary analysis, strong ROR2 protein expression was observed in the DCM-RVF samples with minimal expression in the other groups (**Supplemental Figure 1**). A similarly dramatic relationship was not observed for the other targets; we thus prioritized ROR2 for protein expression analysis in the remaining 123 samples.

ROR2 protein expression in all 143 samples correlated strongly with RNA expression (R^2^ = 0.41, p = 8.1 ×10^−18^). Median and average RVF-to-pRV fold increase in protein expression were 2.0 and 4.5, respectively, (p < 0.05, **Figure 2**). Furthermore, particularly within the RVF group, ROR2 protein expression increased linearly with higher RA:PCWP (R^2^ =0.40, p = 3.0 ×10^−5^, **Figure 2**). Finally, by using the NF ROR2 RNA and protein expression to establish normative ranges, RVF samples demonstrated a greater than 3-fold odds (95^th^ percentile confidence intervals) of expressing ROR2 above the 95^th^ percentile compared to pRV (protein: OR 3.07 (1.1 – 8.4), p = 0.03; RNA: OR 3.18 (1.4 – 7.3), p = 0.0071). In summary, ROR2 expression, both RNA and protein, correlates directly with RVF categorically, and with worse RV hemodynamics.

**Figure 2.**
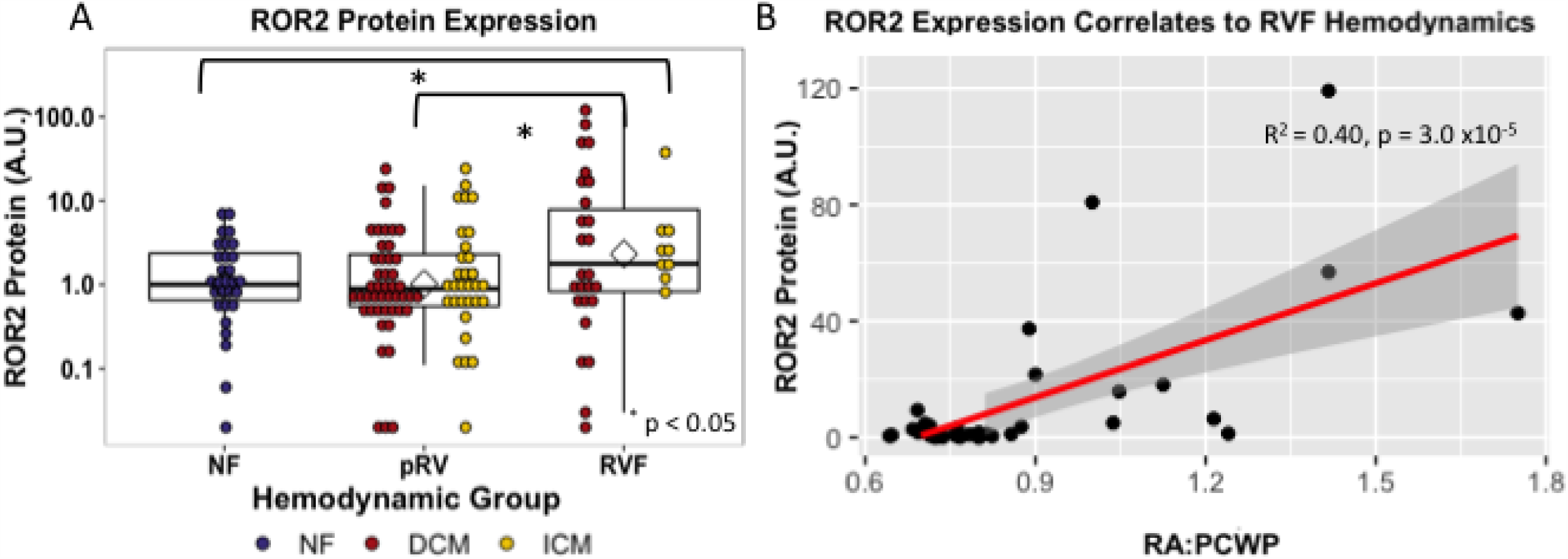
ROR2 protein expression increases in RVF and correlates with hemodynamics. (A) Dot plot demonstrating ROR2 protein expression among different groups and (B) linear regression comparing ROR2 protein expres to RA:PCWP.

#### Impact of ROR2 expression in RVF

##### ROR2 expression correlates with actin cytoskeletal remodeling pathways

We next explored what functional impact higher ROR2 expression might have in the setting of RVF by assessing overall expression and phosphorylation of downstream targets, focusing on cytoskeletal remodeling pathways. Normalized to GAPDH, we found a roughly 1.5 to 2-fold increase in calpain, full length FLNA, and full length spectrin protein expression in high ROR2 expressing RVF patients compared to low ROR2 expression (**Figure 3** and **Table 3**, p < 0.05, for all). Similarly, we identified an almost 8- and 3-fold increase in the calpain cleavage-mediated FLNA (180 kDa) and spectrin (150 kDa) breakdown products, respectively. Phosphorylation of FLNA serine-2152 modulates its mechanosensitive interaction with integrin and inhibits calpain-mediated cleavage, and ROR2 overexpression in HEK293 cells was found to activate one of the known kinases for this site—p21-activated kinase (PAK1)—through a presumed indirect phosphorylation.[33; 34; 35] We therefore assessed the relationship of ROR2 expression, FLNA phosphorylation, and phosphorylated PAK1 expression. In high ROR2 expressing RVF samples, we observed a nearly 10-fold increase in FLNA phosphorylation normalized to total FLNA and a 2-fold increase in phosphorylated PAK1 normalized to GAPDH (p < 0.05, both). Given these consistent relationships observed categorically for high and low ROR2 expression, we next assessed whether expression of these targets correlated linearly or logarithmically with ROR2 expression (**Figure 3** and **Table 3**). The most statistically significant correlation observed was a logarithmic increase in phosphorylated PAK1 with increasing ROR2 expression (R^2^ = 0.83, p = 1.4 × 10^−4^). Full length and phosphorylated FLNA also correlated significantly in a logarithmic relationship with ROR2 expression (R^2^ = 0.67 and 0.62, respectively, p <0.003). Calpain expression correlated linearly with ROR2 expression (R^2^ = 0.66, p = 0.0018). In summary, within the context of RVF, high ROR2 expression correlates with calpain expression and calpain-mediated cleavage as well as an increase in total FLNA, FLNA phosphorylation, and total spectrin expression.

**Table 3.** Comparative analysis of human RVF with high and low ROR2 expression

| Increased ROR2 Expression Correlates with Calpain-Mediated Cleavage in Right Ventricular Failure |  |  |  |  |  |
| --- | --- | --- | --- | --- | --- |
| Target | ROR2 Expression as Continuous Variable |  |  | High vs Low ROR2 Expression |  |
|  | Relationship | Adjusted R<sup>2</sup> | Adj P | Median Fold | Adj P |
| Spectrin 285 kDa | Logarithmic | 0.17 | 0.10 | 1.4 | 0.0080 |
| Spectrin 150 kDa | Logarithmic | 0.17 | 0.10 | 2.7 | 0.015 |
| **FLNA 280 kDa** | **Logarithmic** | **0.67** | **0.0018** | **1.9** | **0.0080** |
| FLNA 190 kDa | Logarithmic | 0.25 | 0.077 | 7.6 | 0.0080 |
| **Phospho:Total FLNA** | **Logarithmic** | **0.62** | **0.0025** | **9.7** | **0.0080** |
| **Phosphorylated PAK1** | **Logarithmic** | **0.83** | **1.4 x10<sup>-4</sup>** | **2.1** | **0.011** |
| **Calpain** | **Linear** | **0.66** | **0.0018** | **1.8** | **0.015** |

4 Comparative analysis of high (n = 6) vs low (n = 6) ROR2 expressing RVF samples using ROR2 expression categorically (high vs 5 low) or as a continuous variable. As a continuous variable, target expression was assessed using both logarithmic and linear models 6 and the most significant model for each is displayed. In bold are comparisons that were statistically significant in both analyses.

**Figure 3.**
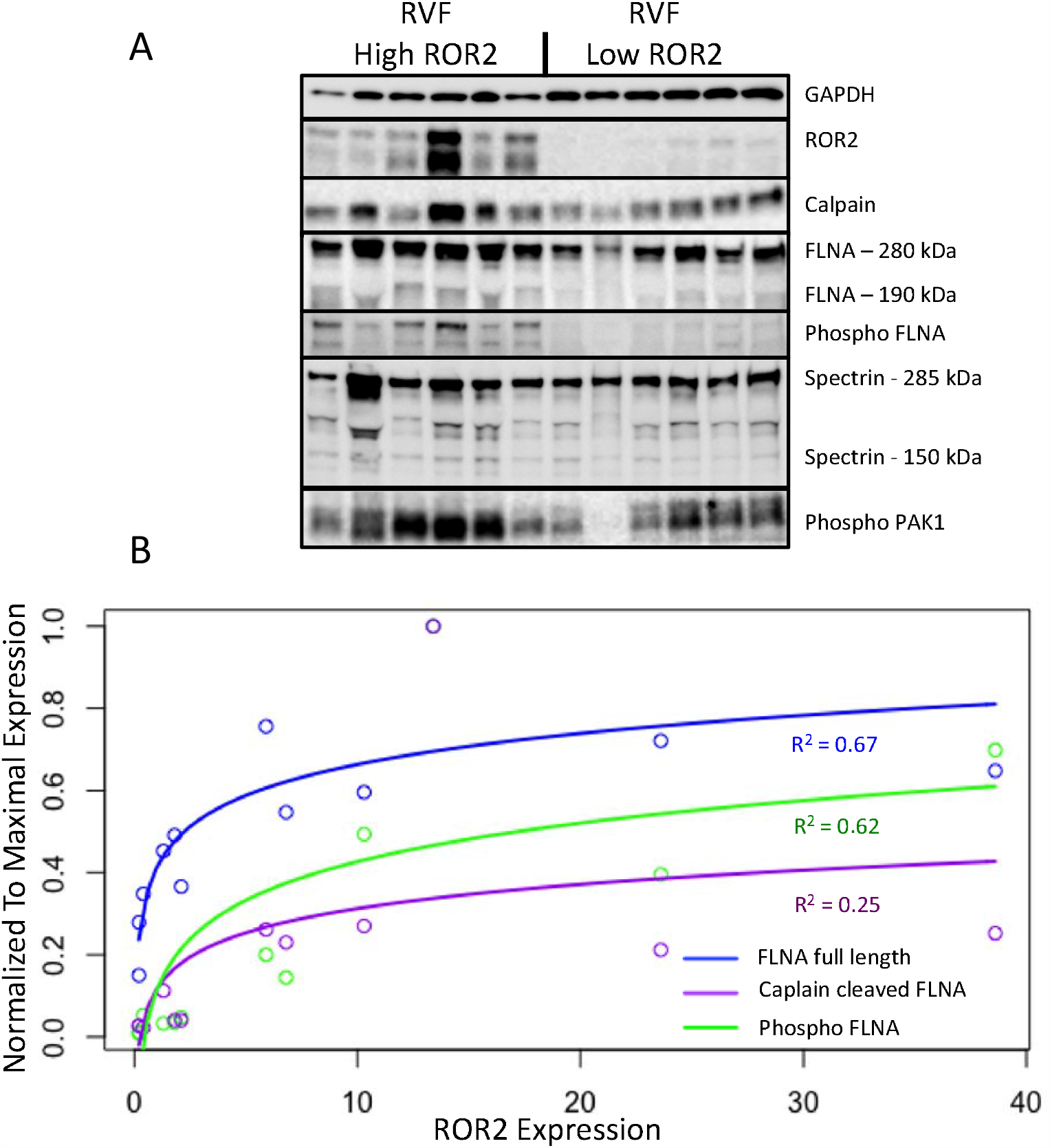
ROR2 expression correlates with increases in calpain expression and calpain mediated cleavage. (A) Western blots comparing expression of downstream targets between high and low ROR2 expressing RVF patients. (B) Logarithmic regressions comparing ROR2 expression to full length, calpain cleaved, and phosphorylated FLNA

## Discussion

To date, there are no evidence-supported therapies that target RVF [9]. Our lack of understanding of the molecular mechanisms that regulate RV remodeling in humans, particularly with respect to adaptive compared to pathologic remodeling, remains a significant barrier towards achieving this goal. Here, we find compelling evidence that non-canonical WNT signaling, and in particular ROR2 signaling, is aberrantly activated in human RVF.

ROR2 is a cell surface receptor tyrosine kinase that transmits noncanonical WNT signaling following binding of its only known ligand—WNT5A—via planar cell polarity, WNT/Ca^2+^, and stress pathways including JNK/cJUN [36]. ROR2 is broadly expressed during embryogenesis, being critical to cardiac, skeletal, and sympathetic nervous system development, but is silent in most healthy postnatal tissue [27; 28; 37]. Global knockout of Ror1 reveals no apparent defects in the developing mouse, but knockout of Ror2 resulted in ventricular septal defects and knockout of both Ror2 and Ror1 led to conotruncal type congenital heart defects [27]. Interestingly, upon our review of their published histology, we noted a significantly noncompacted appearance to the RV and LV myocardium only in the double knockout, which suggests Ror1/2 play a role in myocardial development similar to other noncanonical WNT genes (e.g. disorganized and noncompacted myocardium with loss of *Daam1, Scrib1*, or second heart field-specific loss of *Rac1*) [12; 13; 14]. ROR2 in disease has been largely studied in the context of tumors, where WNT5A/ROR2 signaling leads to activation of calpain-mediated cleavage, cytoskeletal rearrangement for purposes of migration, and as a hypoxia-inducible factor downstream of *VHL/HIF* signaling with implications in tumor invasiveness and metastasis [29; 38; 39]. To date, little is known about the role of ROR2 reactivation in cardiac pathology. In a rat left anterior descending ligation myocardial infarction model, increased protein levels of Wnt5a, Ror2, and Vangl2—one if its immediate downstream targets—were observed in the remote vital area [40]. Using neonatal rat ventricular myocytes (NRVMs), the authors identified phosphorylation of cJun and Jnk as mediators of this activation. Another group found that Wnt5a stimulation of NRVMs caused hypertrophy, cytoskeletal rearrangement, and mPTP opening, but any dependence on Ror2 or other known Wnt5a receptors was not tested [41]. Our results are thus consistent with, and expand on, what few mechanistic data exist on the role of ROR2 in myocardial remodeling, and suggest that ROR2 induction in RVF mechanistically contributes to the pathogenesis of human RVF.

By comparing RVF with high and low ROR2 expression, we found evidence that in the context of RVF ROR2 activation leads to increased calpain-mediated cleavage of FLNA. FLNA is a large scaffolding cytoskeletal protein that is involved in broad cellular functions including maintaining cell and tissue structure (e.g. cross-linking actin filaments, binding integrin, maintaining adherens junctions), cell migration, and as a signaling molecule via nuclear translocation of cleaved fragments [42]. Loss of FLNA in humans or mice causes early lethality with diverse and severe developmental defects affecting the heart, lung, neurologic system, and skeleton [43]. Of note, severe RV myocardial noncompaction has been previously reported in a patient with a *FLNA* G1728C mutation [44]. FLNA has been connected to ROR2 in cells other than cardiomyocytes. For example, in melanoma cells, ROR2-FLNA interaction is critical for WNT5A-induced JNK phosphorylation, cytoskeleton remodeling, and cell migration, and WNT5A-ROR2 binding induces calpain expression and its cleavage of FLNA leading to increased invasiveness [29; 45]. Importantly, there is also evidence that calpain contributes to pathologic RV remodeling. In an acute pressure overload-induced rabbit RVF model, direct right coronary artery infusion of the selective calpain inhibitor MDL-28710 partially rescued RV function and reduced cleavage of the cytoskeletal protein talin [46]. Thus, our findings of ROR2 induction and activation of its downstream pathways, suggest a model (**Figure 4**) whereby an embryonic WNT5a/ROR2 program is reactivated in the setting of RVF, which then promotes calpain-mediated cleavage of FLNA and likely other targets, ultimately leading to maladaptive cytoskeletal changes and worsening RVF.

**Figure 4.**
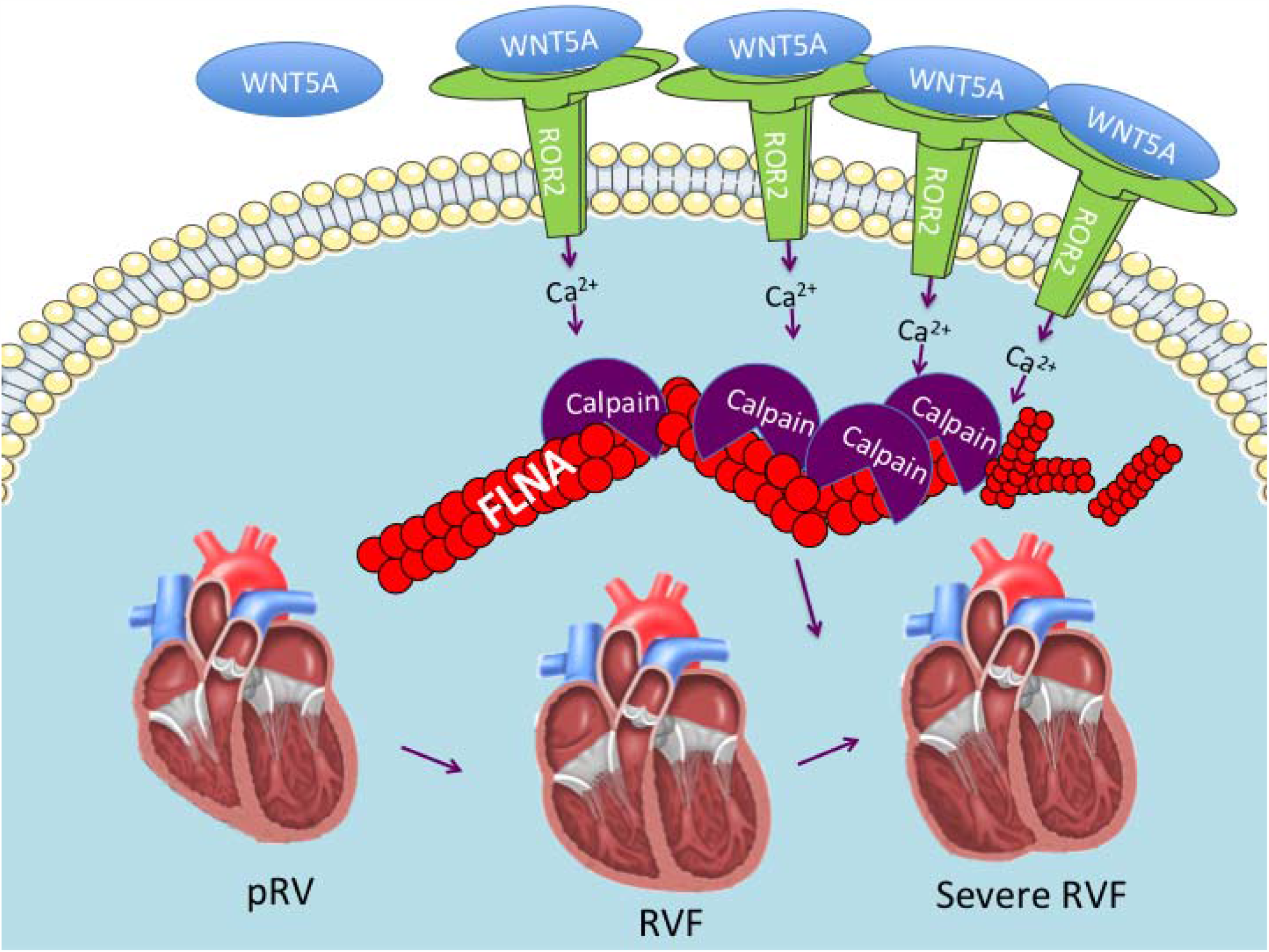
Proposed RVF model consisting of reactivation of a ROR2 fetal gene program with increasing RVF severity, which results in increased calpain expression and activity leading to calpain-mediated cleavage of cytoskeleton structural proteins including FLNA.

## Limitations

There may have been some disease progression or clinical deterioration in the time interval between tissue collection and hemodynamic assessments for DCM and ICM RV, although if so, such changes would likely have introduced variability and weakened our differential expression analyses. It is also possible that changes in gene expression occurred during the period between heart harvest and tissue freezing, although this process is highly controlled by the Penn Human Heart Tissue Library with standard operating procedures including assiduous adherence to cold cardioplegia and snap freezing samples in liquid nitrogen. Our use of rare human-derived tissue allowed us to examine differential expression in a natural setting, but it did limit our ability to mechanistically evaluate the impact of altered ROR2 expression. Thus, further work will be necessary such as testing the response to RV pressure or volume overload in Ror2 gain-of-function and loss-of-function mice.

## Conclusions

In this study, using one of the largest and most diverse existing libraries of human heart tissue, we found that WNT signaling is broadly dysregulated in RV remodeling in the setting of ICM and DCM. We found a large reactivation of embryonic ROR2 RNA and protein expression in RVF, which correlated with worse RV hemodynamics, and with activation of a downstream pathway of increased calpain expression and FLNA cleavage. Taken together, the data reveal the robust activation of noncanonical WNT signaling in human RVF, and identify ROR2 as a potential novel RVF therapeutic target that may suppress cytoskeletal remodeling.

## Data Availability

The datasets generated and analyzed for the current study are available upon reasonable request to the corresponding author.

## Data Availability Statement

All datasets generated and analyzed for the current study are available in supplementary material.

## Ethics Statement

Procurement of all RV myocardial tissue was performed using Gift-of-Life and University of Pennsylvania Institutional Review Board (approval 802781) approved protocols with informed consent provided when appropriate. The authors attest they are in compliance with human studies committees of the authors’ institutions and Food and Drug Administration guidelines, including patient consent where appropriate.

## Author contributions

Conceptualization and Design (JJE, KBM, DB, SR, ZPA). Acquisition of data (JJE, JB, LL, DQH, KCB, DM, HTH, MZ). Analysis and interpretation of data (JJE, JB, SJ, NY, SW, SR, ZPA). Writing and editing of manuscript (JJE, SJ, KBM, SR, ZPA).

## Conflicts of Interest

The authors have no relationships relevant to the content of this paper to disclose.

## Acknowledgements

The authors acknowledge all patients and donors who provided tissue without which this study could not have been performed. The authors acknowledge the Gift-of-Life Donor Program Philadelphia, PA who helped provide access to nonfailing tissue from unused donor hearts. A previous version of this manuscript was submitted to the preprint server medRxiv (https://doi.org/10.1101/2020.07.03.20134965).

## Sources of Funding

This study was supported by grants from NIH and National Heart, Lung, and Blood Institute (NHLBI) 152446 (to Z.PA.), K08 HL127277-01 (to S.R.), and R01 HL105993 (to K.B.M), and Department of Defense W81XWH18-1-0503 (to Z.P.A.) and PR151448 (to DB and SR). JJE was supported by NIH 5T32HL007915, Matthew’s Heart of Hope, and Congenital Heart Disease Coalition.

## Non-standard Abbreviations

CST: Cell Signaling Technology
DCM: dilated cardiomyopathy
FLNA: filamin A
ICM: ischemic cardiomyopathy
LVF: left ventricular failure
NF: nonfailing
PAK1: p-21 activated kinase
pRV: preserved right ventricular function
RA: right atrial pressure
RA:PCWP: right atrial to pulmonary capillary wedge pressure ratio RT-PCR real time polymerase chain reaction
RV: right ventricle
RVF: right ventricular failure

## SUPPLEMENTAL MATERIAL

**Supplemental Table 1.**
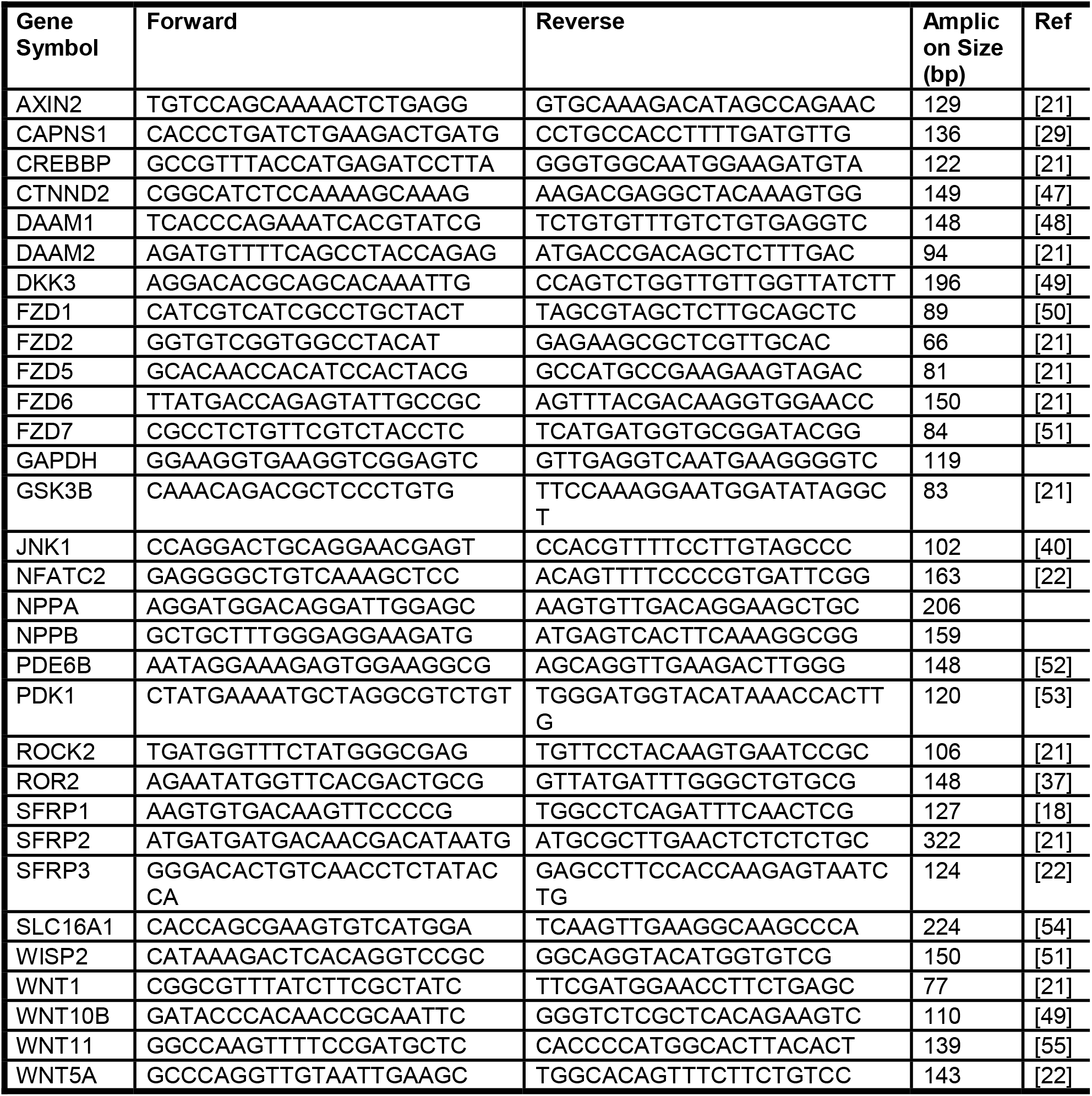
RT-PCR primer sequences for tested genes with references to known role in WNT signaling and/or cardiac pathology.

**Supplemental Table 2.** Clinical and demographic characteristics of combined DCM and ICM hemodynamic groups.

| Clinical and Demographic Characteristics |  |  |  | 4 |
| --- | --- | --- | --- | --- |
| Variables | RVF<br>n = 35 | pRV<br>n = 78 | Adj p |  |
| DCM | 74% | 60% | 0.77 |  |
| Age<sup>a</sup> | 57<br>(52, 61) | 55.5<br>(49, 62) | 0.92 |  |
| Male | 60% | 78% | 0.37 |  |
| Ethnicity |  |  | 0.88 |  |
| Caucasian | 63% | 72% |  |  |
| African American | 29% | 23% |  |  |
| Other | 9% | 5% |  |  |
| Heart weight (grams)<sup>a</sup> | 499<br>(388, 570) | 498<br>(429, 609) | 0.87 |  |
| Weight (kg)<sup>a</sup> | 84<br>(73, 97) | 81<br>(70, 93) | 0.88 |  |
| Body surface area (m<sup>2</sup>)<sup>a</sup> | 1.96<br>(1.9, 2.2) | 1.97<br>(1.8, 2.2) | 0.88 |  |
| GFR<sup>#</sup> | 50.1<br>(40, 62) | 62.2<br>(49, 81) | 0.16 |  |
| Diabetes Mellitus | 38% | 32% | 0.89 |  |
| Insulin | 23% | 16% | 0.88 |  |
| Thyroid medication | 31% | 10% | 0.16 |  |
| Pacer | 31% | 15% | 0.37 |  |
| ACE inhibitor | 54% | 55% | 1.0 |  |
| ARB | 29% | 22% | 0.88 |  |
| β-blocker | 77% | 92% | 0.36 |  |
| Any Remodeling | 100% | 96% | 0.88 |  |
| Calcium Channel Blocker | 6% | 4% | 1.0 |  |
| Digoxin | 43% | 46% | 1.0 |  |
| Diuretic | 94% | 90% | 0.89 |  |
| Lipid lowering | 49% | 59% | 0.88 |  |
| Milrinone | 70% | 73% | 1.0 |  |

<sup>a</sup>Continuous variables presented as median (interquartile range) and *p* value using two-tailed Mann-Whitney U test. Categorical variables presented as percent and *p* value using Chi square contingency tables using pairwise deletion for any missing data. All *p* values are Benjamini-Hochberg corrected, and none were less than 0.05. Abbreviations: angiotensin converting enzyme inhibitor (ACE inhibitor), angiotensin receptor blocker (ARB), coronary artery bypass grafting (CABG), glomerular filtration rate (GFR), and RCA (right coronary artery disease).

**Supplemental Table 3.** Demographic characteristics of NF compared to RV remodeling groups.

| Demographic Characteristics Between Nonfailing and RV Remodeling Groups |  |  |  |  |  |  |  |
| --- | --- | --- | --- | --- | --- | --- | --- |
| Variables | NF<br>n = 29 | DCM-pRV<br>n= 47 | DCM-RVF<br>n= 26 | Adj P | ICM-pRV<br>n = 31 | ICM-RVF<br>n= 9 | Adj P |
| Age<sup>a</sup> | 56<br>(45, 63) | 52<br>(44, 60) | 56.5<br>(48, 58) | 0.34 | 61.0<br>(55, 63) | 60.0<br>(56, 62) | 0.56 |
| Male | **38%** | **72%** | **50%** | **0.047** | **84%** | **89%** | **4.6 x 10<sup>-4</sup>** |
| Ethnicity |  |  |  | 0.34 |  |  | 0.69 |
| Caucasian | 79% | 64% | 54% |  | 78% | 89% |  |
| African American | 14% | 30% | 38% |  | 13% | 0% |  |
| Other | 7% | 6% | 8% |  | 3% | 11% |  |
| Weight (kg)<sup>a</sup> | 69<br>(62, 87) | 80<br>(69, 92) | 76<br>(73, 91) | 0.83 | 82<br>(75, 94) | 93<br>(84, 113) | 0.56 |
| Body surface area (m<sup>2</sup>)<sup>a</sup> | 1.82<br>(1.7, 2.0) | 1.95<br>(1.8, 2.1) | 1.90<br>(1.8, 2.1) | 0.30 | 1.99<br>(1.9, 2.2) | 2.19<br>(2.0, 2.4) | 0.41 |

<sup>a</sup>Continuous variables presented as median (interquartile range) comparing NF/pRV/RVF separately for DCM and ICM using Kruskal-Wallis test. Gender and ethnicity assessed using chi-square. All comparisons corrected using Benjamini-Hochberg

**Supplemental Table 4.**
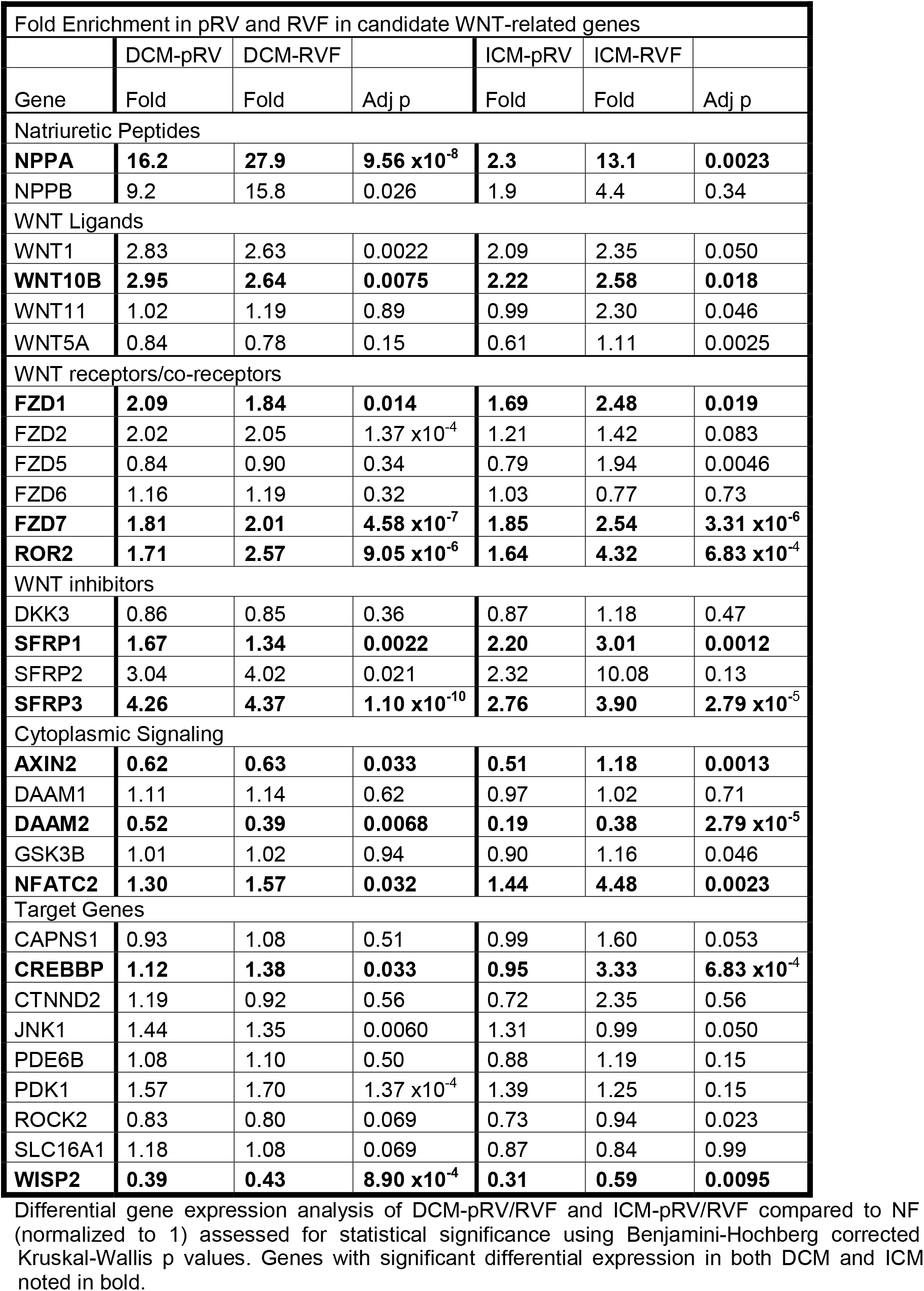
Differential WNT-related gene expression in Dilated and Ischemic Cardiomyopathy Right Ventricle

| Fold Enrichment in pRV and RVF in candidate WNT-related genes |  |  |  |  |  |  |
| --- | --- | --- | --- | --- | --- | --- |
|  | DCM-pRV | DCM-RVF |  | ICM-pRV | ICM-RVF |  |
| Gene | Fold | Fold | Adj p | Fold | Fold | Adj p |
| Natriuretic Peptides |  |  |  |  |  |  |
| **NPPA** | **16.2** | **27.9** | **9.56 x10<sup>-8</sup>** | **2.3** | **13.1** | **0.0023** |
| NPPB | 9.2 | 15.8 | 0.026 | 1.9 | 4.4 | 0.34 |
| WNT Ligands |  |  |  |  |  |  |
| WNT1 | 2.83 | 2.63 | 0.0022 | 2.09 | 2.35 | 0.050 |
| **WNT10B** | **2.95** | **2.64** | **0.0075** | **2.22** | **2.58** | **0.018** |
| WNT11 | 1.02 | 1.19 | 0.89 | 0.99 | 2.30 | 0.046 |
| WNT5A | 0.84 | 0.78 | 0.15 | 0.61 | 1.11 | 0.0025 |
| WNT receptors/co-receptors |  |  |  |  |  |  |
| **FZD1** | **2.09** | **1.84** | **0.014** | **1.69** | **2.48** | **0.019** |
| FZD2 | 2.02 | 2.05 | 1.37 x10<sup>-4</sup> | 1.21 | 1.42 | 0.083 |
| FZD5 | 0.84 | 0.90 | 0.34 | 0.79 | 1.94 | 0.0046 |
| FZD6 | 1.16 | 1.19 | 0.32 | 1.03 | 0.77 | 0.73 |
| **FZD7** | **1.81** | **2.01** | **4.58 x10<sup>-7</sup>** | **1.85** | **2.54** | **3.31 x10<sup>-6</sup>** |
| **ROR2** | **1.71** | **2.57** | **9.05 x10<sup>-6</sup>** | **1.64** | **4.32** | **6.83 x10<sup>-4</sup>** |
| WNT inhibitors |  |  |  |  |  |  |
| DKK3 | 0.86 | 0.85 | 0.36 | 0.87 | 1.18 | 0.47 |
| **SFRP1** | **1.67** | **1.34** | **0.0022** | **2.20** | **3.01** | **0.0012** |
| SFRP2 | 3.04 | 4.02 | 0.021 | 2.32 | 10.08 | 0.13 |
| **SFRP3** | **4.26** | **4.37** | **1.10 x10<sup>-10</sup>** | **2.76** | **3.90** | **2.79 x10<sup>-5</sup>** |
| Cytoplasmic Signaling |  |  |  |  |  |  |
| **AXIN2** | **0.62** | **0.63** | **0.033** | **0.51** | **1.18** | **0.0013** |
| DAAM1 | 1.11 | 1.14 | 0.62 | 0.97 | 1.02 | 0.71 |
| **DAAM2** | **0.52** | **0.39** | **0.0068** | **0.19** | **0.38** | **2.79 x10<sup>-5</sup>** |
| GSK3B | 1.01 | 1.02 | 0.94 | 0.90 | 1.16 | 0.046 |
| **NFATC2** | **1.30** | **1.57** | **0.032** | **1.44** | **4.48** | **0.0023** |
| Target Genes |  |  |  |  |  |  |
| CAPNS1 | 0.93 | 1.08 | 0.51 | 0.99 | 1.60 | 0.053 |
| **CREBBP** | **1.12** | **1.38** | **0.033** | **0.95** | **3.33** | **6.83 x10<sup>-4</sup>** |
| CTNND2 | 1.19 | 0.92 | 0.56 | 0.72 | 2.35 | 0.56 |
| JNK1 | 1.44 | 1.35 | 0.0060 | 1.31 | 0.99 | 0.050 |
| PDE6B | 1.08 | 1.10 | 0.50 | 0.88 | 1.19 | 0.15 |
| PDK1 | 1.57 | 1.70 | 1.37 x10<sup>-4</sup> | 1.39 | 1.25 | 0.15 |
| ROCK2 | 0.83 | 0.80 | 0.069 | 0.73 | 0.94 | 0.023 |
| SLC16A1 | 1.18 | 1.08 | 0.069 | 0.87 | 0.84 | 0.99 |
| **WISP2** | **0.39** | **0.43** | **8.90 x10<sup>-4</sup>** | **0.31** | **0.59** | **0.0095** |

Differential gene expression analysis of DCM-pRV/RVF and ICM-pRV/RVF compared to NF (normalized to 1) assessed for statistical significance using Benjamini-Hochberg corrected
- 1 Kruskal-Wallis p values. Genes with significant differential expression in both DCM and ICM - 2 noted in bold.

**Supplemental Figure 1.**
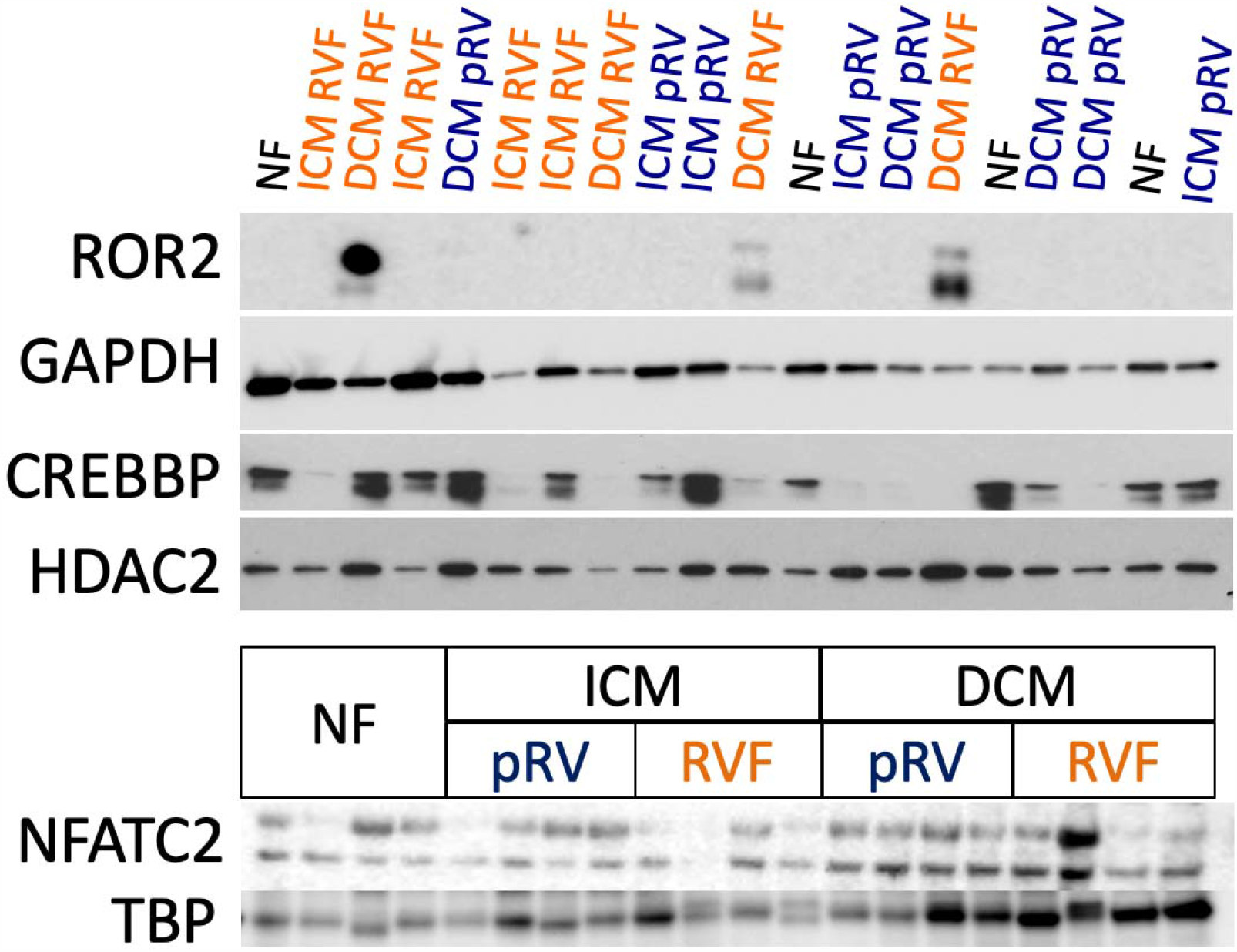
Preliminary western blot analysis comparing four representative samples from each group using the highest and lowest RA:PCWP for RVF and pRV, respectively, and the lowest NPPA expression for NF to represent the extremes of each group.

## Notes

### Competing Interest Statement

The authors have declared no competing interest.

### Author Declarations

Procurement of all myocardial tissue was performed using Gift-of-Life and University of Pennsylvania Institutional Review Board (approval 802781) approved protocols with informed consent provided when appropriate.

